# Schooling amidst a pandemic: parents’ perceptions about reopening schools and anticipated challenges during COVID-19

**DOI:** 10.1101/2021.03.02.21252777

**Authors:** Ankita Meghani, Smisha Agarwal, Alexander John Zapf, Jeffrey G. Edwards, Alain Labrique, Dustin Gibson

**Affiliations:** Johns Hopkins Bloomberg School of Public Health, Department of International Health, Baltimore, MD, United States

**Author notes:** Corresponding author’s address: Johns Hopkins Bloomberg School of Public Health, 615 N Wolfe St, Baltimore, MD 21205.

## Abstract

**Introduction:** During the COVID-19 pandemic, numerous states in the United States instituted measures to close schools or shift them to virtual platforms. Understanding parents’ preferences for sending their children back to school, and their experiences with distance learning is critical for informing school reopening guidelines. This study characterizes parents’ plans to send their children to school, and examines the challenges associated with school closures during the 2020-2021 academic year.

**Methods:** A national-level cross-sectional online survey was conducted in September 2020. Focusing on a subset of 510 respondents, who were parents of school-aged children, we examined variations in parents’ plans for their children to return to school by their demographic and family characteristics, and challenges they anticipated during the school-year using multivariable logistic regressions.

**Results:** Fifty percent of respondents (n=249) said that they would send their children back to school, 18% (n=92) stated it would depend on what the district plans for school reopening, and 32% (n=160) would not send their children back to school. No demographic characteristics were significantly associated with parents plans to not return their children to school. Overall, parents reported high-level of access to digital technology to support their child’s learning needs (84%). However, those who reported challenges with distance learning due to a lack of childcare were less likely not to return their children to school (aOR=0.33, 95% CI: 0.17, 0.64). Parents who reported requiring supervision after school had higher odds of having plans to not return their children to school (aOR=1.97, 95% CI: 1.03, 3.79). Parents viewed COVID-19 vaccines and face masks important for resuming in-person classes.

**Discussion:** About one-third of parents surveyed objected to their children returning to school despite facing challenges with distance learning. Besides access to vaccines and face masks, our findings highlight the need to better equip parents to support remote learning, and childcare.

## Introduction

In a public health effort to decrease transmission of severe acute respiratory syndrome-coronavirus-2 (SARS-CoV-2), which causes coronavirus disease (COVID-19), several states in the United States instituted measures to close schools or shift them to virtual platforms starting in March 2020 [1]. Most states made the decision to close schools in April 2020 given the challenges of maintaining safe distances, the potential for transmission in a closed school environment, and the risks, especially to teachers and staff, many of whom may constitute a high risk group due to preexisting health conditions and older age [2]. Given the significant impact school closures can have on children’s learning, and associated economic impact on their parents [3], the necessity and appropriate implementation of school closures as well measures for safe reopening need to be continuously assessed.

School administrators are grappling with issues of adequate ventilation in classrooms, physical distancing, and implementing proper sanitation of playground equipment, doorknobs, and other frequently used surfaces to prevent potential transmission of SARS-CoV-2 [4]. The average classrooms in the United States contain more than 20 students, requiring the implementation of staggered school schedules and other strategies to promote physical distancing [5]. Decisions around school closures and reopening also impact teachers and educators who are balancing the trade-offs between in-person teaching and increased risk of COVID-19 transmission [6]. Likewise, employers are contending with the effects of school closures/reopening; having children in school allows parents to return to work, enabling businesses and communities to function more effectively.

Across all stakeholders, the greatest impact of school closures has perhaps been felt by parents of school-aged children. Many parents struggle with supporting their children’s remote learning needs while meeting work responsibilities [7]. In particular, women who tend to shoulder work, and the bulk of childcare and other caregiving responsibilities, have been most affected by the pandemic [8]. The burden associated with school closures has disproportionately impacted parents, who lack job flexibility [9–11], have little or no childcare options [12,13], and have inadequate access to educational and technology resources to support remote schooling. School closures pose further challenges to parents whose children rely on school for meals [14–16], access to counseling [17–19], and other extra-curricular resources necessary to support healthy child development [20]. For many parents, who are struggling to cope with their children’s educational and development needs at home, school reopening may be a priority.

Poorer households may not have access to necessary equipment such as personal computers and high speed internet to facilitate remote schooling [21]. Such challenges may be greater in households with several school-aged children each of whom may need attention and personal computers. Additionally, parents who earn daily wages or do not have the ability to work from home may need to leave their children at home without adequate oversight to support their education, which can have long-term consequences for the child’s learning. School closures also disproportionately affect children who rely on schools feeing programs, and whose families may be poorer, have lower levels of education, and have access to limited resources [22]. Parents in these positions may be more inclined to send their children back to school to avail these benefits. The impact of COVID-19 has been known to disproportionately impact African American and Hispanic parents, a significant portion of the essential workforce, who may have limited flexibility to work from home, or provide the necessary support required for distance learning [23,24].

Parents with different household characteristics and other socio-demographic factors may express different concerns around remote learning and school reopening. In light of recent evidence suggesting similar rates of transmission among older children and adolescents, and high levels of viral load among children under five years of age [25], multigenerational households, for example, living with grandparents, or households with older parents or parents with chronic health conditions may be reluctant to resume in-person instruction compared to households with younger parents and children. Specifically, emerging evidence suggests the new variant of SARS-CoV-2 has a higher propensity to infect children [26]. Even parents with children of different age groups (teenagers versus primary school-aged children) may have different concerns. For example, parents of older children, who may be required to provide less oversight, may be less inclined to resume schooling compared to parents of a primary school-aged child, who would likely benefit significantly from the pedagogy provided by a schoolteacher.

Understanding parents’ preferences for sending their children back to school, and their experiences, with remote learning is critical for informing school reopening guidelines. In this study, we strive to: (i) characterize, parents’ willingness to send their children to school, overall and by demographic characteristics, and (ii) examine challenges associated with school closures by parents’ demographics. These findings may guide school districts in making informed decisions between resuming in-person instruction and maintaining remote/virtual instruction.

## Methods

### Study Design

This study was nested within the broader ‘Pandemic Pulse’ survey developed by researchers at the Johns Hopkins Bloomberg School of Public Health to monitor the impact of the COVID-19 pandemic in the United States. A national cross-sectional online survey was conducted from September 1^st^ to September 7^th^, 2020. Dynata (https://www.dynata.com), an online platform that maintains a database of over 62 million unique users who have provided their demographic information [27], was contracted to administer surveys to users based on a set of pre-defined demographic characteristics. Dynata randomly selected US residents from its database and emailed online surveys to eligible participants. The study sample was matched to the 2019 United States Census population estimates for age, race, gender, income, and census region. Dynata implements several quality control procedures to ensure high data quality, such as implementation of digital fingerprinting to prevent duplication of respondents and verification of respondent profiles Participants over 18 years of age were eligible to participate in the survey. Those who completed the survey received a small financial incentive from Dynata.

### Survey tool

The survey tool comprised of questions relating to nine thematic modules: 1) demographics, 2) risk perceptions, 3) education, 4) viral testing, 5) stigma and agency, 6) COVID-19 knowledge, 7) medication and treatment perceptions, 8) pregnancy and antenatal care and 9) vaccine perceptions and information. This study presents the findings from the education module, which asked respondents who identified as parents about: 1) their plan to have their children return to school during the 2020-2021 school year; 2) challenges they face or anticipate facing with meeting their child’s educational needs during the school year, including access to technology to support remote learning; and 3) important considerations for school reopening. Survey questions are provided in **S1 Table**. These questions were adapted from a Washington County Public School Survey [28].

### Sample size

The sample size for the overall study (N=1592) was calculated to produce nationally representative estimates with a margin of error of +/-3%. To be able to detect 10% differences in indicators across White, African American, and Latino racial/ethnic groups, the sample for African American and Latino respondents was each increased to 385 participants. This study focuses on a subset of 510 survey respondents, who self-reported being parents of school-aged children, including kindergartners.

### Measures

#### Outcomes

The primary outcome of this study was whether parents planned for their child to return to school in the 2020-2021 school year. Parents were asked “At this time, are you planning to return your child to school for the 2020-2021 school year?” There were three response options: (1) Yes, (2) No, and (3) Depends on what plans the school district makes. The primary outcome was recategorized into a binary variable: (i) “no”; and (ii) “possible plans to return” (combining “yes” and “it depends on school district plans” categories) for further analyses.

#### Parents’ demographic and family characteristics

Parents’ age, race, education, income, the school-level of children in the family, and the type of school they attend were collected in the survey. Age was assessed in six categories (18-24; 25-34; 35-44; 45-54; 55-64; and 65+ years). Race/ethnicity were queried according to six categories (American Indian or Alaskan Native; Asian or Pacific Islander; Black or African American; Hispanic or Latino; White/Caucasian; and other. Income level was measured by asking about the current annual household income from all sources in: (i) $10,000 increments for up to $50,000; (ii) $50,000 – $69,000; (iii) $70,000 – $84,999; (iv) 85,000 – 99,999; (v) increments of $50,000 for $100,000 – $199,999 and (vi) $200,000 and above. Education was assessed in six categories (less than high school degree; high school degree or equivalent (e.g., GED); some college but no degree; associate degree; bachelor’s degree; and graduate degree).

#### Challenges parents anticipate facing during the 2020-2021 school year

Parents were asked to anticipate challenges with meeting their child’s educational needs during the school year. Parents responded to a “select all that apply” question that asked “Which of the following statements are true when thinking of the 2020-2021 school year? Respondents could choose one or more from seven options: (1) My child cannot carry out distance learning from home due to lack of child care; (2) Carrying out distance learning from home will place an extremely difficult burden on my family; (3) My child will require supervision at school before the school day begins; (4) My child will require supervision at school after the school day ends; (5) My child will rely on the school/district for meals and receiving enough food; (6) My child will rely on school district transportation (school bus); and (7) None of these apply to me. In addition, parents were asked about their access to necessary technology resources (e.g. laptop, high-speed internet) to support remote learning by answering: (1) yes; (2) no; or (3) maybe.

#### Factors influencing parents’ plans to return their children to school

Parents were asked to select five out of the fourteen factors (e.g., COVID-19 vaccine availability, staggered scheduled, temperature screening, face mask use) listed as priorities important to consider when schools reopen for in-person classes. The response options are listed in **S1 Table**.

### Statistical analysis

Survey data were weighted by race and census region using 2019 Census estimates, given that Blacks and Hispanics were oversampled. We performed descriptive statistics and examined the frequency distributions of the two outcomes by demographic characteristics, including age, race, education, income, the school level of children in the family, and the type of school they attend. Bivariate relationships were conducted using Pearson’s chi-square tests between parents’ demographic characteristics and questions about parents’ plans for their child returning to school (objective 1), challenges parents’ face with meeting their child/children’s educational needs during the school year (objective 2), and factors influencing parents’ plans to return their children to school (objective 3).

We estimated unadjusted and adjusted odds ratios using multivariable logistic regressions to examine differences in the primary outcome, parents’ plan about their child/children not returning to school during the 2020-2021 school year by parents’ demographic characteristics. To assess whether the anticipated challenges parents face in the 2020-2021 school year were associated with their plans for their children to return to school, we also used multivariable logistic regressions controlling for parents’ demographic characteristics.

All analyses were conducted in Stata 14 (StataCorp LLC, College Station, TX) [29]. The SVY package, with subpopulations option, was used for all analyses.

### Ethical approval

The study protocol and survey instruments were approved by the Institutional Review Board at Johns Hopkins Bloomberg School of Public Health (IRB00012413). All participants provided electronic consent at the beginning of the survey.

## Results

### Sample characteristics

Sample characteristics of parent participants, and their weighted responses are presented in **Table 1**. The sample included 510 participants with 72% (n=367) between 25-44 years of age, with roughly an even distribution by gender (53% female, n= 268). African American (n=126) and Latino (n=134) respondents each represented approximately one fourth of this sample. More than 50% of participants had at least a bachelor’s or graduate degree (n=267). Roughly 25% (n=124) were earning $40,000 to $69000, 22% (n=111) were earning $70,000 to $99,999 and 27% (n=138) were earning more than $100,000 per annum. Sixty eight percent of participants (n=342) reported that their children attend public schools. Eleven percent (n=58) of participants had children in day-care, nearly half had children in elementary and middle school, 22% in high school (n= 111), and 19% with children attending more than one type of school (n= 95).

**Table 1.** Demographic and socioeconomic characteristics for respondents who self-reported being parents of kindergarteners and school-aged children (N=510)

|  | **Unweighted**<br>**n (%)** | **Weighted<sup>a</sup>**<br>**%** |
| --- | --- | --- |
| **Age** |  |  |
| 18-24 | 44 (8.63) | 9.55 |
| 25-34 | 196 (38.43) | 34.85 |
| 35-44 | 171 (33.53) | 36.54 |
| 45-54 | 71 (13.92) | 13.88 |
| 55+ | 28 (5.49) | 5.18 |
| **Gender** |  |  |
| Female | 268 (52.76) | 52.06 |
| Male | 240 (47.24) | 47.94 |
| **Race** |  |  |
| White | 215 (42.16) | 58.31 |
| African American | 126 (24.71) | 11.40 |
| Latino | 134 (26.27) | 19.37 |
| Other<sup>#</sup> | 35 (6.86) | 10.91 |
| **Education** |  |  |
| High School or less | 87 (17.09) | 16.28 |
| Associate degree | 70 (13.75) | 12.20 |
| Some college but no degree | 85 (16.70) | 16.99 |
| Bachelors | 154 (30.26) | 30.49 |
| Graduate | 113 (22.2) | 24.04 |
| **Income** |  |  |
| <\$20,000 | 58 (11.39) | 9.47 |
| \$20,000 to \$39,999 | 78 (15.32) | 14.09 |
| \$40,000 to \$69,999 | 124 (24.36) | 22.91 |
| \$70,000 to \$99,999 | 111 (21.81) | 23.94 |
| ≥\$100,000 | 138 (27.11) | 29.59 |
| **School level of child** |  |  |
| Daycare | 58 (11.37) | 10.08 |
| Elementary & Middle | 246 (48.24) | 51.33 |
| High School | 111 (21.76) | 21.37 |
| More than one school type | 95 (18.63) | 17.23 |
| **School type of child** |  |  |
| Public | 342 (68.13) | 73.58 |
| Private or religious | 160 (31.87) | 26.4 |

<sup>a</sup>Weighted data for race by Census region, <sup>#</sup>Includes American Indians, Asians and other racial/ethnic groups. Noted all the categories may not add up to 510 because some respondents refused to answer the question.

### Parents’ plans to return their children to school (Objective 1)

Of the 501 participants who responded to this question (**S2 Table**), fifty percent of participants (n=249) said that they would send their children back to school, and 18% (n=92) stated it would depend on what the district plans for school reopening, while 32% (n=160) would not send their children back to school. Respondents decisions did not significantly vary by the type of school their child attended (public or private; p=0.236) and school level (daycare, elementary, middle, high school; p=0.160). Differences in views on school reopening were also not statistically different by race (p=0.136), education (p=0.088), income-levels (p=0.182). However, female participants were significantly more likely to report not sending their children back to school (59% female vs. 42% male, p-value = 0.011).

After controlling for other demographic variables, Latino/Hispanic ethnicity was associated with higher odds of parents not planning for their child to return to school during the 2020-2021 school year, but was not statistically significant (aOR=1.68, 95% CI: 0.99, 2.84, p=0.053) (**Table 2**). Gender, education, income, school level of child, and the type of school attended by the child were not significantly associated with parents’ plans for not returning their child to school.

**Table 2.** Results from multivariable logistic regression models estimating unadjusted and adjusted odds ratio for demographic characteristics associated with parents’ plans to not have their child return to school

|  | Unadjusted odds<br>(95% CI) | p-value | Adjusted Odds Ratio<br>(95% CI) | p-value |
| --- | --- | --- | --- | --- |
| **Age** |  |  |  |  |
| 18-24 | Reference |  | Reference |  |
| 25-34 | 1.60 (0.64, 3.94) | 0.306 | 1.49 (0.53 , 4.17) | 0.441 |
| 35-44 | 1.72 (0.67, 4.39) | 0.252 | 1.98 (0.68 , 5.75) | 0.204 |
| 45-54 | 1.95 (0.71, 5.30) | 0.190 | 2.14 (0.70 , 6.52) | 0.179 |
| 55+ | 0.91 (0.26, 3.21) | 0.892 | 1.03 (0.26 , 4.00) | 0.960 |
| **Gender** |  |  |  |  |
| Female | Reference |  | Reference |  |
| Male | 0.75 (0.48, 1.18) | 0.225 | 0.80 (0.50 , 1.27) | 0.356 |
| **Race** |  |  |  |  |
| White | Reference |  | Reference |  |
| African American | 1.20 (0.73, 1.96) | 0.459 | 1.33 (0.77 , 2.29) | 0.291 |
| Latino | 1.42 (0.88, 2.30) | 0.142 | 1.68 (0.99 , 2.84) | 0.053 |
| Other | 0.50 (0.13, 1.84) | 0.299 | 0.63 (0.17 , 2.34) | 0.496 |
| **Education** |  |  |  |  |
| High School or less | Reference |  | Reference |  |
| Associate degree | 0.92 (0.44, 1.92) | 0.833 | 0.81 (0.37 , 1.75) | 0.596 |
| Some college but no degree | 1.26 (0.62, 2.59) | 0.512 | 1.14 (0.52 , 2.45) | 0.736 |
| Bachelors | 0.59 (0.30, 1.16) | 0.130 | 0.56 (0.27 , 1.16) | 0.125 |
| Graduate | 0.69 (0.36, 1.35) | 0.286 | 0.57 (0.26 , 1.23) | 0.157 |
| **Income** |  |  |  |  |
| <\$20,000 | Reference | | Reference | |
| \$20,000 to \$39,999 | 0.61 (0.26, 1.44) | 0.264 | 0.70 (0.29 , 1.71) | 0.443 |
| \$40,000 to \$69,999 | 0.66 (0.31, 1.41) | 0.291 | 0.87 (0.39 , 1.91) | 0.737 |
| \$70,000 to \$99,999 | 0.46 (0.20, 1.03) | 0.061 | 0.75 (0.32 , 1.74) | 0.504 |
| >\$100,000 | 0.58 (0.27, 1.21) | 0.149 | 0.95 (0.40 , 2.21) | 0.909 |
| **School level of child** |  |  |  |  |
| Daycare | Reference |  | Reference |  |
| Elementary & Middle | 0.67 (0.34, 1.33) | 0.262 | 0.76 (0.32 , 1.79) | 0.538 |
| High School | 0.59 (0.27, 1.27) | 0.180 | 0.61 (0.23 , 1.59) | 0.316 |
| More than one school type | 0.46 (0.21, 1.02) | 0.058 | 0.44 (0.16 , 1.17) | 0.102 |
| **School type of child** |  |  |  |  |
| Public | Reference |  | Reference |  |
| Private or religious | 1.10 (0.68, 1.78) | 0.685 | 1.22 (0.65 , 2.28) | 0.525 |

### Challenges parents anticipate with meeting their children’s educational needs and their influence on parents’ plans to return their children to school (Objective 2)

Eighty-four percent (n=427) of all parents had access to technology (e.g., laptop or high-speed internet) for remote learning, with 13% (n=64) reporting they did not have access, and 3% (n=15) reporting no need to use technology (**S3 Table**). Access to technology did not significantly vary by parents’ age, gender, race, education level, and income level. However, access to technology for remote learning was significantly associated with the type of school the child attended (p=0.004), and the child’s school level (p=0.005).

**Fig 1** summarizes these challenges parents anticipated facing during the 2020-2021 school year. Fifty-eight percent (n=296) of parents anticipated challenges related to distance learning if the school moved to a remote format. Thirty-seven percent (n=188) of parents required child supervision at school either before the school day begins or after the school day ends with 15% (n=79) of parents indicating their reliance on schools for their children’s meals and 12% (n=59) for transportation.

**Fig 1.**
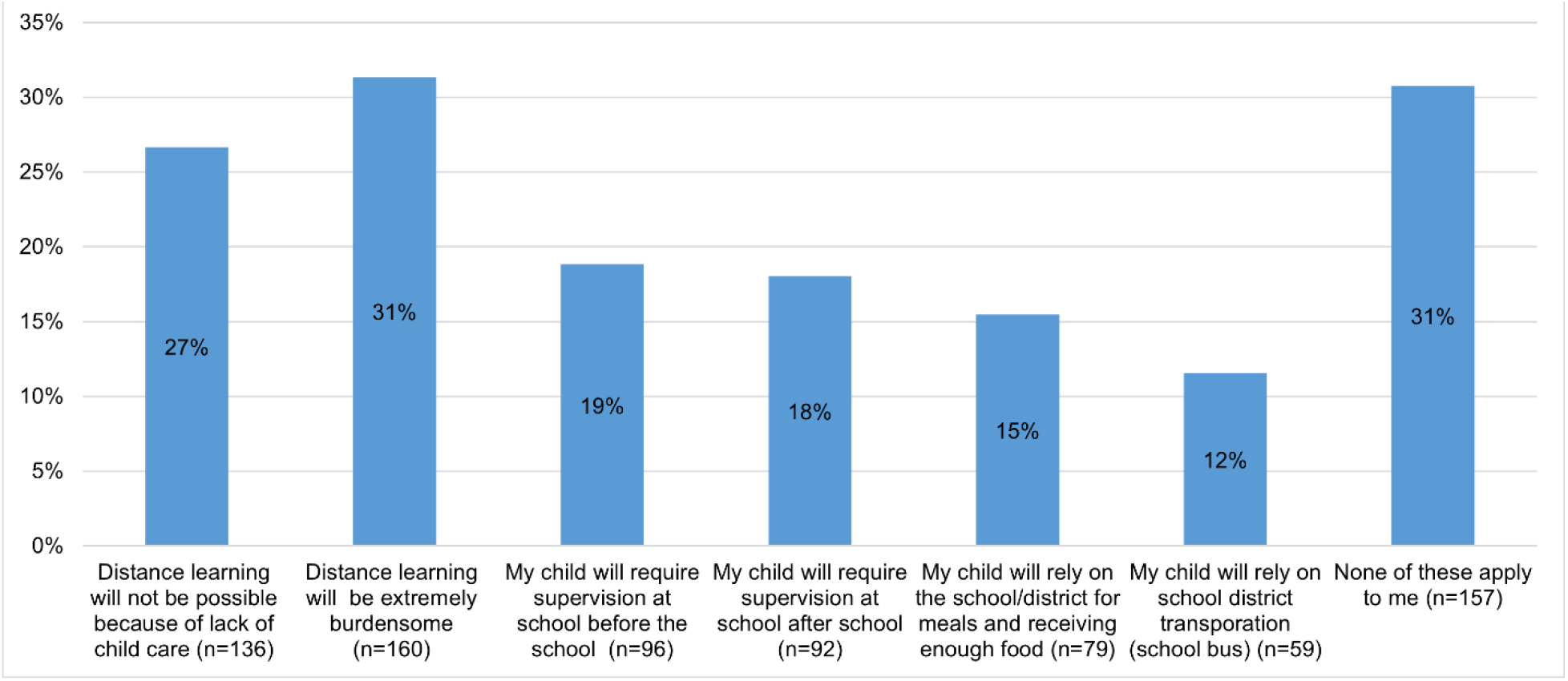
Challenges parents anticipate facing during the 2020-2021 school year. Note: Cumulative percentage could be greater than 100% because respondents were allowed to select multiple options in responding to this question about challenges they anticipated during the 2020-2021 school year.

**Table 3** shows the relationship between each challenge parents anticipated during the school year (exposure) with their plans to not have their child return to school (outcome). After controlling for sociodemographic characteristics in the adjusted models, parents who reported challenges with distance learning due to a lack of childcare had lower odds of having plans to not return their children to school (aOR=0.33, 95% CI: 0.17, 0.64). Conversely, parents who reported requiring supervision after school in the evening had higher odds of having plans to not return their children to school (aOR=1.97, 95% CI: 1.03, 3.79). Four challenges - distance learning being viewed as a burden, supervision of children before school, reliance on school for meals, and reliance on school district for transportation - were not significantly associated with parents’ plans to not return their children to school.

**Table 3.**
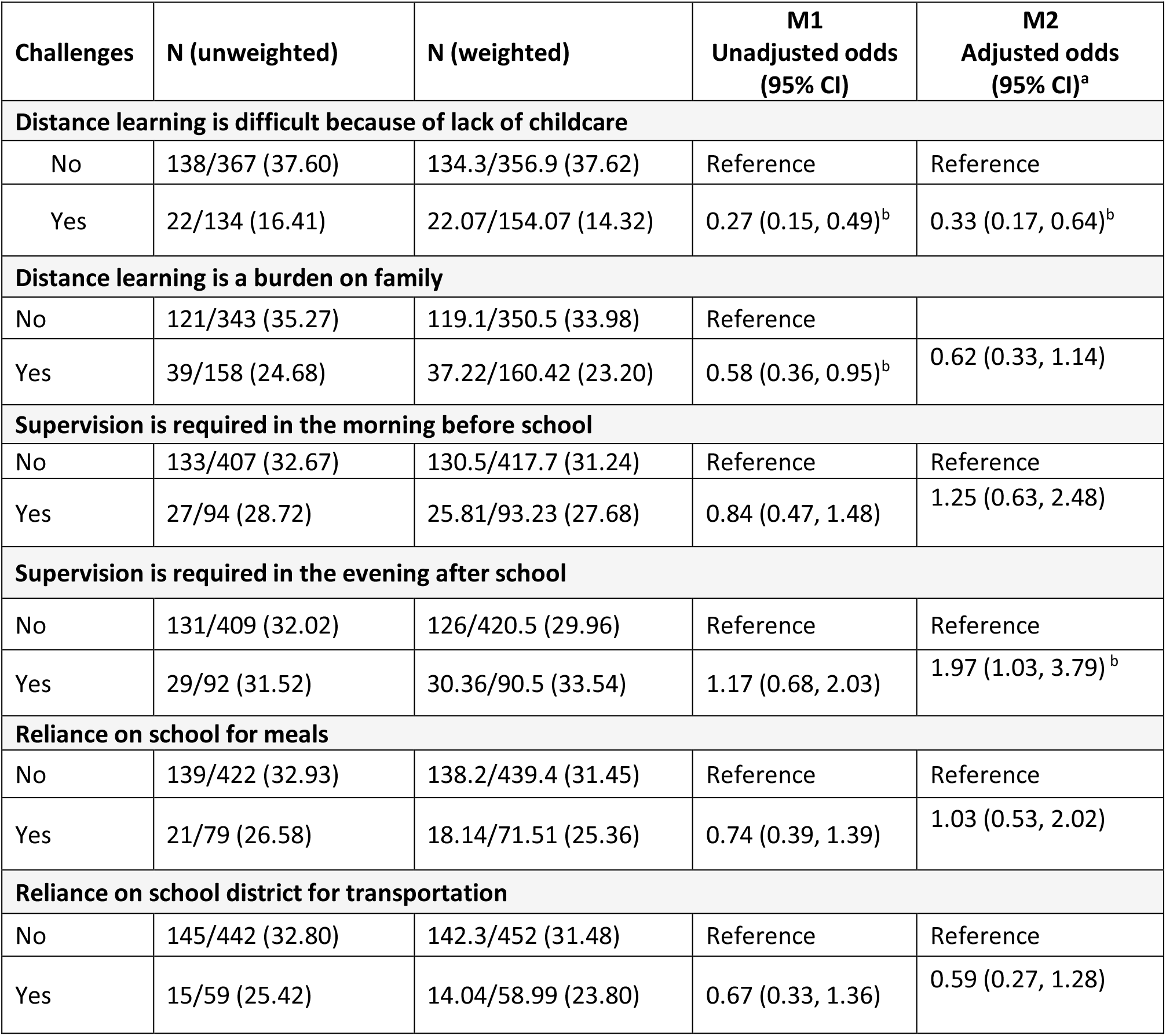
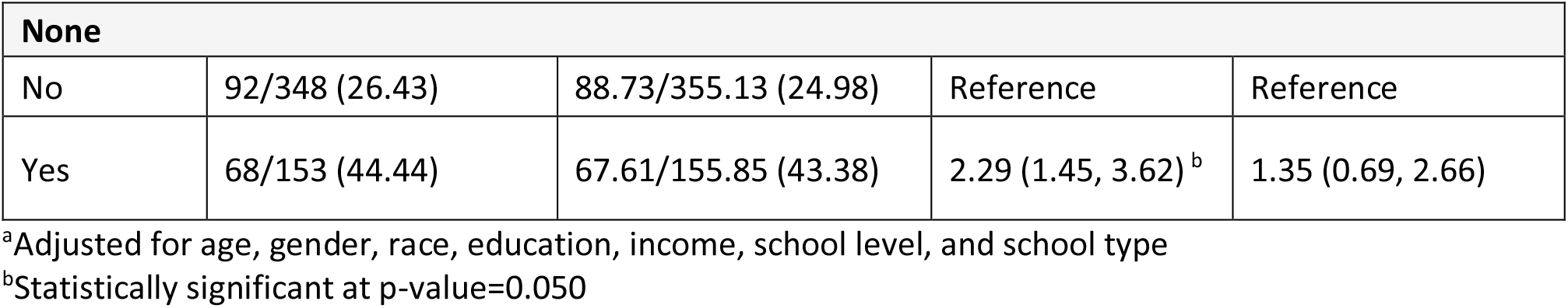
Results from unadjusted logistic regression models (M1) compared to adjusted multivariable models (M2), including 95% confidence intervals (CI).

### Factors influencing parents’ plans to return their children to school (Objective 3)

**Fig 2** summarizes factors influencing parents’ plans to have their children return to school. Among these, parents said they would only return their children to school if a vaccine was available (29%, n=145), and safety guidelines (29%, n=145) for social distancing, hand washing, use of face coverings and temperature checks were encourage. Another 18% (n=89) of parents said their child would only return their children to school if the safety guidelines were enforced (see **Fig 2**).

**Fig 2.**
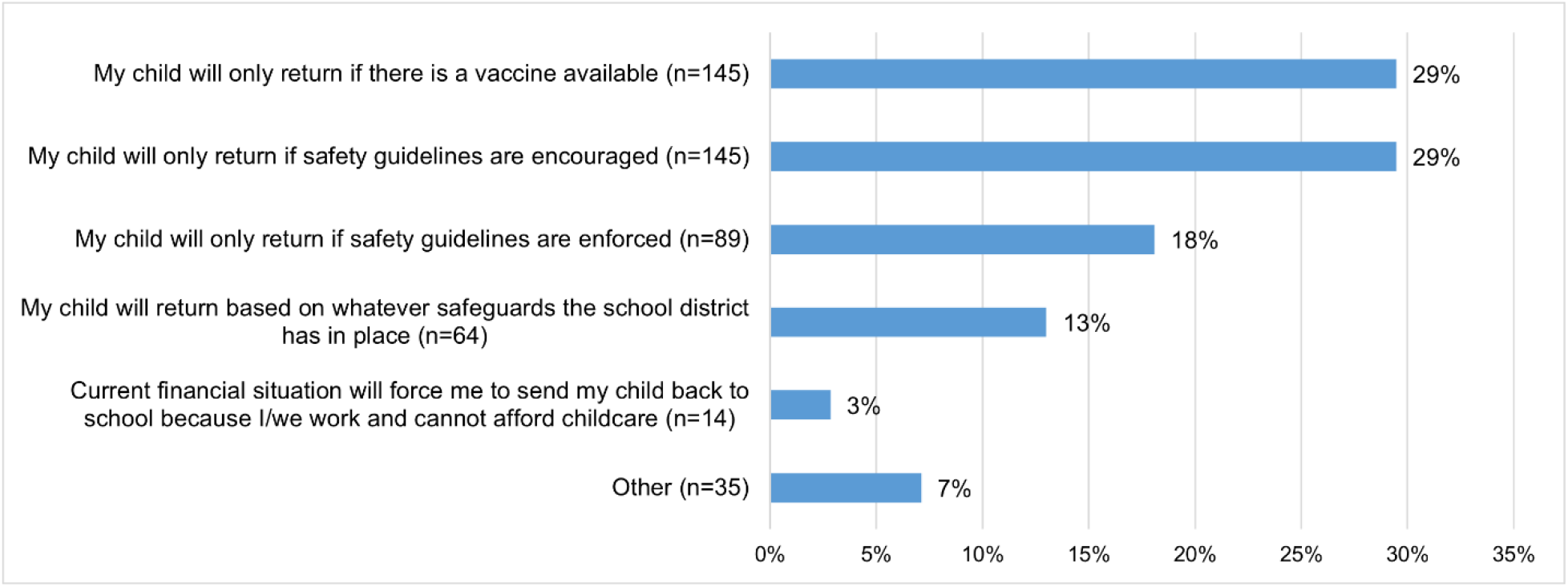
Factors influencing parents’ plans to have their children return to school (n=492) Note: Safety guidelines refer to social distancing, handwashing, face-covering guidelines, and temperature checks.

When asked to select the top five priorities for school reopening from a list (**Table 4**), parents viewed COVID-19 vaccine availability (n=184) as a top priority followed by staff being required to wear a mask or face covering (n=182) and masks being available for everyone (n=171) and students being required to wear a mask (n=170). COVID-19 testing and COVID-19 antibody testing availability (n=140), hand sanitizer availability in class and its use (n=116) and temperature screening for students and staff each day (n=114) were also identified as the remaining priorities.

**Table 4.** Parents that selected one of the below factors as the top 5 priorities for reopening schools and resuming in-person classes.

| Factors to consider for reopening schools and resuming in-person classes | n |
| --- | --- |
| COVID-19 vaccine availability | 184 |
| Requirement for teachers/staff to wear a mask/face-covering | 182 |
| Availability of masks/face-coverings for everyone | 171 |
| Requirement for students to wear a mask/face-covering | 170 |
| COVID-19 testing and COVID-19 antibody testing availability | 140 |
| Hand sanitizer is provided in each classroom and is frequently used | 116 |
| Temperature screening for students and staff each day | 114 |
| Staff are trained on CDC guidance to reduce likelihood of COVID-19 transmission | 93 |
| Limiting classroom seating to maintain social distancing | 91 |
| Regularly scheduled, adult supervised hand-washing | 87 |
| Enhanced cleaning of surfaces in the schools. | 76 |
| Staggering schedules to reduce crowding | 60 |
| Cafeteria seating is spaced for proper social distancing and food in individually packaged | 57 |
| Other | 23 |

Note: Respondents were asked to select their top five priorities from the list of 14 factors.

## Discussion

In helping states determine how to reopen schools safely, understanding parents’ perspectives on school reopening, and challenges they are currently facing or anticipating in meeting their children’s distance learning needs is important. Our findings show that 32% of parents would not have their children return to school during the 2020-2021 school year. Among the socio-demographic characteristics assessed in this study, only Latino/Hispanic ethnicity of parents was associated with borderline statistically significant higher odds of planning to have their children not return to school.

A priori, we had expected parents in poorer households to report greater challenges with access to equipment for distance learning, such as personal computers and high-speed internet. Our findings, however, showed there were no significant differences in access by sociodemographic characteristics, including race/ethnicity, income, and education levels. Consistent with other findings that found high-level of technology access to support home schooling [30], our study found that 84% of parents had access to digital technology to support their child’s learning needs.

Despite these high-levels of access, we did observe variations in technology access by school type of children with those in public schools having greater access to technology than those in private schools. This may be explained by increased distribution of tablets and computers by rural and low-income school districts during the pandemic to bridge a potential digital divide [31]. Though about 59% of teachers surveyed by Rand Corporation reported that nearly all or most of their students have access to the necessary technology for remote learning, they also reiterated that inadequate access to high-speed internet and technology platforms would significantly affect remote learning [32]. It is important that households’ access to technology is routinely monitored because it may be a critical driver in potential disparities that may affects students’ learning during the pandemic.

Our analysis also found that parents who reported challenges associated with distance learning were more likely to have their children return to school. For many parents, challenges with distance learning have been attributed to the difficulty of balancing their work-related responsibilities with caring for their children and families [30]. Recent studies examining the mental health impact of school closures have shown declines in parents mental well-being, with nearly a third of parents reporting a worsening of mental health and close to 40% of parents demonstrating signs of depression or anxiety, respectively [19,33,34].

Parents also face difficult decisions regarding their children’s health and education, which requires consideration of a wide array of factors to reassure parents that sending their children back to school is safe. For example, special precautions may be required for children with underlying health conditions that predispose them to increased disease risk or children living in households with individuals at increased risk for severe disease, e.g., those with medical conditions and older adults [35].

The struggle with distance learning for parents has been exacerbated with the recent acceleration of COVID-19 transmission in communities across the United States during the winter months, which has resulted in the reimplementation of school closures in many states and counties [36–38]. Against the backdrop of nationwide surges and continuing uncertainty about the trajectory of the pandemic in 2021, decision-makers are faced with balancing complex trade-offs between reducing disease spread on the population level and negative effects on the economy as well as detrimental impacts on the development and wellbeing of children [39,40].

To support parent’s decision-making about sending their children back to school, they should be provided with accurate information about their school’s safety or risk mitigation practices [41]. School districts in collaboration with their local health departments are having to decide on parameters (e.g., community infection rates) for reopening their schools. However, it is unclear how parents’ preferences are being incorporated in these decisions. Our study found parents expressed preference for safety measures - such as, the use of masks, as well as availability of COVID-19 vaccines, COVID-19 testing and hand sanitizers - that should be implemented within schools. It is important that these safety measures be enforced (not just recommended) so that parents feel more reassured about sending their children back to school in a safe manner.

### Limitations

The findings of this study should be interpreted in the light of some limitations. Survey participants were drawn from the Dynata database and were not randomly drawn from the general population, potentially limiting the generalizability of our findings. Furthermore, participating in the survey requires access to a device with internet access. While smartphone ownership has been increasing, and the digital divide has narrowed in the United States [42], it is possible our online survey may not captured the experiences of individuals without the resources and technology to access the internet [43], which may be associated with lower levels of education and low-income households [44]. In addition, we did not collect demographic data about parents’ underlying health risks, such as chronic health diseases, and whether respondents belonged to intergenerational households, which may have also influenced their responses.

Despite our large sample size for the overall survey, the substrata for specific age groups and racial or ethnic minorities were relatively small, which may have reduced the statistical precision of our estimates. While our study systematically oversampled African American and Hispanic racial/ethnic groups, our estimates for smaller racial/ethical groups are not generalizable for parents. Finally, residual confounding may limit our estimates as we did not capture data on potential confounders, such as living in multigenerational households, employment of the parent (e.g., as an essential worker), and medical conditions among family members.

## Conclusion

This study underscores that nearly one-third of parents in the United States object to returning their children to school during the 2020-21 school-year despite facing significant challenges. Families need to be supported by federal, state, or local governments to protect them against potential detrimental losses in their children’s education and other long-term impacts. Besides safe and effective vaccines and face masks to mitigate transmission of COVID-19, our findings highlight the need to provide adequate resources for remote learning, and to support families meet their childcare needs.

## Data Availability

Data are available upon request.

## Supporting information

**S1 Table**. Survey questions included in this analysis.

**S2 Table**. Parents’ plans to return their children to school in the 2020-2021 academic year.

**S3 Table**. Parents’ access to technology for remote learning during the 2020-2021 academic year.

