## Supplementary Tables for "Schooling amidst a pandemic: parents’ perceptions about reopening schools and anticipated challenges during COVID-19"

**S1 Table.** Survey questions included in this analysis

| | |
|---|---|
| At this time, are you planning to return your child to school for the 2020-2021 school year | 1=Yes<br>0=No<br>2=Unsure<br>3=It depends on what plans the school district makes<br>4=Prefer not to say |
| Which of the following statements are true when thinking of the 2020-2021 school year? Select all that apply | 1= My child cannot carry out distance learning from home due to lack of child care<br>2= Carrying out distance learning from home will place an extremely difficult burden on my family<br>3= My child will require supervision at school before the school day begins<br>4= My child will require supervision at school after the school day ends<br>5= My child will rely on the school/district for meals and receiving enough food<br>6= My child will rely on school district transportation (school bus)<br>7= None of these apply to me<br>8= Prefer not to say |
| Do you have access to necessary technology resources (e.g., Laptop, High-speed internet) to support your child's remote learning, if required ? | 1=Yes<br>2=No<br>3=Didn't need to use any technology<br>98=Prefer not to say |
| Which of the following statements best describes your overall feeling about returning your child to school? | 1 = My child will only return if there is a vaccine available<br>2 = My child will only return if social distancing, handwashing, face-covering guidelines, and temperature checks are encouraged<br>3 = My child will only return if social distancing, handwashing, face-covering guidelines, and temperature checks are enforced<br>4 = My current financial situation will force me to send my child back to school because I/we work and cannot afford childcare<br>5 = My child will return based on whatever safeguards the school district has in place<br>6 = Prefer not to say<br>7 = Other |
| Many factors are important to consider when we reopen schools. Please select your **top five** priorities regarding health safety practices if schools were to open for in-person classes this fall | 1=COVID-19 vaccine availability<br>2=COVID-19 testing and COVID-19 antibody testing availability<br>3=Availability of masks/face-coverings for everyone<br>4= Requirement for teachers/staff to wear a mask/face-covering<br>5= Requirement for students to wear a mask/face-covering<br>6= Regularly scheduled, adult supervised hand-washing<br>7= Staff are trained on CDC guidance to reduce likelihood of COVID-19 transmission<br>8= Temperature screening for students and staff each day<br>9=Hand sanitizer is provided in each classroom and is frequently used<br>10=Cafeteria seating is spaced for proper social distancing and food in individually packaged<br>11= Staggering schedules to reduce crowding |

| | |
|---|---|
| | 12= Limiting classroom seating to maintain social distancing<br>13= Enhanced cleaning of surfaces in the schools.<br>14= Other |

**S2 Table.** Parents' plans to return their children to school in the 2020-2021 academic year

| | Parents plans to return their children to school during the 2020-2021 academic year | | | | |
|---|---|---|---|---|---|
| | Yes N (%) | No N (%) | Depends[a] N (%) | Total N (%) | p-value |
| **Age** | | | | | |
| 18-24 | 25 (10.04) | 10 (6.25) | 7 (7.61) | 42 (8.38) | |
| 25-34 | 101 (40.56) | 60 (37.50) | 31 (33.70) | 192 (38.32) | |
| 35-44 | 82 (32.93) | 59 (36.88) | 28 (30.43) | 169 (33.73) | |
| 45-54 | 29 (11.65) | 24 (15.00) | 17 (18.48) | 70 (13.97) | |
| 55+ | 12 (4.82) | 7 (4.38) | 9 (9.78) | 28 (5.59) | 0.307 |
| **Gender** | | | | | |
| Female | 113 (45.56) | 95 (59.38) | 53 (58.24) | 261 (52.30) | |
| Male | 135 (54.44) | 65 (40.63) | 38 (41.76) | 238 (47.70) | 0.011 |
| **Race** | | | | | |
| White | 115 (46.18) | 64 (40.00) | 33 (35.87) | 212 (42.32) | |
| African American | 58 (23.29) | 40 (25.00) | 25 (27.17) | 123 (24.55) | |
| Latino | 54 (21.69) | 48 (30.00) | 30 (32.61) | 132 (26.35) | |
| Other | 22 (8.84) | 8 (5.00) | 4 (4.35) | 34 (6.79) | 0.136 |
| **Education** | | | | | |
| HS or less | 36 (14.46) | 32 (20.00) | 18 (19.78) | 86 (17.20) | |
| Associate degree | 27 (10.84) | 24 (15.00) | 18 (19.78) | 69 (13.80) | |
| Some college but no degree | 37 (14.86) | 31 (19.38) | 13 (14.29) | 81 (16.20) | |
| Bachelors | 89 (35.74) | 38 (23.75) | 26 (28.57) | 153 (30.60) | |
| Graduate | 60 (24.10) | 35 (21.88) | 16 (17.58) | 111 (22.20) | 0.088 |
| **Income** | | | | | |
| <$20,000 | 25 (10.04) | 22 (13.84) | 9 (9.78) | 56 (11.20) | |
| $20,000 to $39,999 | 36 (14.46) | 24 (15.09) | 17 (18.48) | 77 (15.40) | |
| $40,000 to $69,999 | 49 (19.68) | 39 (24.53) | 30 (32.61) | 118 (23.60) | |
| $70,000 to $99,999 | 63 (25.30) | 32 (20.13) | 16 (17.39) | 111 (22.20) | |
| >100K+ | 76 (30.52) | 42 (26.42) | 20 (21.74) | 138 (27.60) | 0.182 |
| **School level of child** | | | | | |
| Daycare | 25 (10.04) | 22 (13.75) | 10 (10.87) | 57 (11.38) | |
| Elementary & Middle | 114 (45.78) | 83 (51.88) | 42 (45.65) | 239 (47.70) | |
| High School | 53 (21.29) | 30 (18.75) | 27 (29.35) | 110 (21.96) | |
| More than one school type | 57 (22.89) | 25 (15.63) | 13 (14.13) | 95 (18.96) | 0.160 |
| **School type of child** | | | | | |
| Public | 160 (65.31) | 106 (67.95) | 69 (75.00) | 335 (76.83) | |
| Private or religious | 85 (34.69) | 50 (32.05) | 23 (25.00) | 101(23.17) | 0.236 |

[a]Depends on what plans the district makes

**S3 Table.** Parents' access to technology for remote learning during the 2020-2021 academic year

| | Access to tech for remote learning | | | |
|---|---|---|---|---|
| | Yes<br>n (%) | No<br>n (%) | Don't need to use<br>n (%) | p-value |
| **Age** | | | | |
| 18-24 | 37 (8.67) | 4 (6.25) | 2 (13.33) | |
| 25-34 | 154 (36.07) | 31 (48.44) | 9 (60.00) | |
| 35-44 | 152 (35.6) | 17 (26.56) | 2 (13.33) | |
| 45-54 | 64 (14.99) | 6 (9.38) | 1 (6.67) | |
| 55+ | 20 (4.68) | 6 (9.38) | 1 (6.67) | 0.135 |
| **Gender** | | | | |
| Female | 228 (53.4) | 30 (47.62) | 8 (53.33) | |
| Male | 199 (46.6) | 33 (52.38) | 7 (46.67) | 0.692 |
| **Race** | | | | |
| White | 180 (42.15) | 27 (42.19) | 7 (46.67) | |
| African American | 105 (24.59) | 18 (28.13) | 3 (20.00) | |
| Latino | 114 (26.70) | 13 (20.31) | 4 (26.67) | |
| Other | 28 (6.56) | 6 (9.38) | 1 (6.67) | 0.918 |
| **Education** | | | | |
| HS or less | 77 (18.03) | 8 (12.50) | 2 (13.33) | |
| Associate degree | 59 (13.82) | 10 (15.63) | 1 (6.67) | |
| Some college but no degree | 70 (16.39) | 8 (12.5) | 5 (33.33) | |
| Bachelors | 129 (30.21) | 20 (31.25) | 4 (26.67) | |
| Graduate | 92 (21.55) | 18 (28.13) | 3 (20.00) | 0.620 |
| **Income** | | | | |
| <$20,000 | 49 (11.50) | 9 (14.06) | 0 (0.00) | |
| $20,000 to $39,999 | 65 (15.26) | 8 (12.50) | 4 (26.67) | |
| $40,000 to $69,999 | 100 (23.47) | 15 (23.44) | 7 (46.67) | |
| $70,000 to $99,999 | 98 (23.00) | 12 (18.75) | 0 (0.00) | |
| >100K+ | 114 (26.76) | 20 (31.25) | 4 (26.67) | 0.192 |
| **School level of child** | | | | |
| Daycare | 40 (9.37) | 11 (17.19) | 6 (40.00) | |
| Elementary & Middle | 208 (48.71) | 29 (45.31) | 6 (40.00) | |
| High School | 100 (23.42) | 10 (15.63) | 1 (6.67) | |
| More than one school type | 79 (18.50) | 14 (21.88) | 2 (13.33) | 0.005[a] |
| **School type of child** | | | | |
| Public | 299 (71.19) | 34 (53.97) | 7 (46.67) | |
| Private or religious | 121 (28.81) | 29 (46.03) | 8 (53.33) | 0.004[a] |

[a]statistically significant at p-value=0.050
